# Translating Deep Learning to Clinical Practice: External Validation and Clinical Benefit of an Electrocardiogram-Based Neural Network for Detecting Low Ejection Fraction

**DOI:** 10.1101/2025.10.02.25336997

**Authors:** Nicole Hakim, Greg Lee, David M. Vidmar, Loren P. Wagner, Matthew Trachtenberg, Christopher M. Haggerty, Brandon K. Fornwalt, John Pfeifer

## Abstract

Low ejection fraction (EF), an indicator of impaired heart function, often goes undiagnosed and can lead to avoidable heart failure and arrhythmias. We developed and externally validated a deep learning model for detecting low EF from 12-lead electrocardiograms. The model achieved 85.8% sensitivity and 83.0% specificity on an independent validation cohort, with consistent results across demographic subgroups. These findings supported FDA 510(k) clearance of the model. Clinical net benefit analysis further showed that the model provides greater clinical value than default screening approaches, confirming its meaningful potential impact for clinical practice.

## 1 Introduction

Left ventricular ejection fraction (EF) is a critical marker of cardiac function, with EF*≤* 40% indicating increased risk for heart failure and arrhythmias. Many cases go undiagnosed because symptoms are non-specific and echocardiography (echo), the standard clinical diagnostic, requires specialized staff and resources. This makes routine screening difficult in many settings. While electrocardiograms (ECGs) are inexpensive and widely used, clinicians cannot reliably detect low EF from ECGs alone. Recent advances in machine learning have shown that deep neural networks can identify complex patterns in ECG data that are not apparent to clinicians, enabling scalable screening for low EF (1; 2). However, most prior work has lacked large-scale, diverse, external validation (3).

In this work, we developed and externally validated a deep learning model for low EF detection from 12-lead ECGs, using a multi-institutional dataset and an independent cohort of over 16,000 patients from four US health systems. The model achieved high sensitivity and specificity across demographic subgroups, supporting its FDA 510(k) clearance. We further assessed clinical utility using net benefit analysis, demonstrating that the model provides greater value than default screening strategies and is well-suited for real-world deployment.

**Figure 1.**
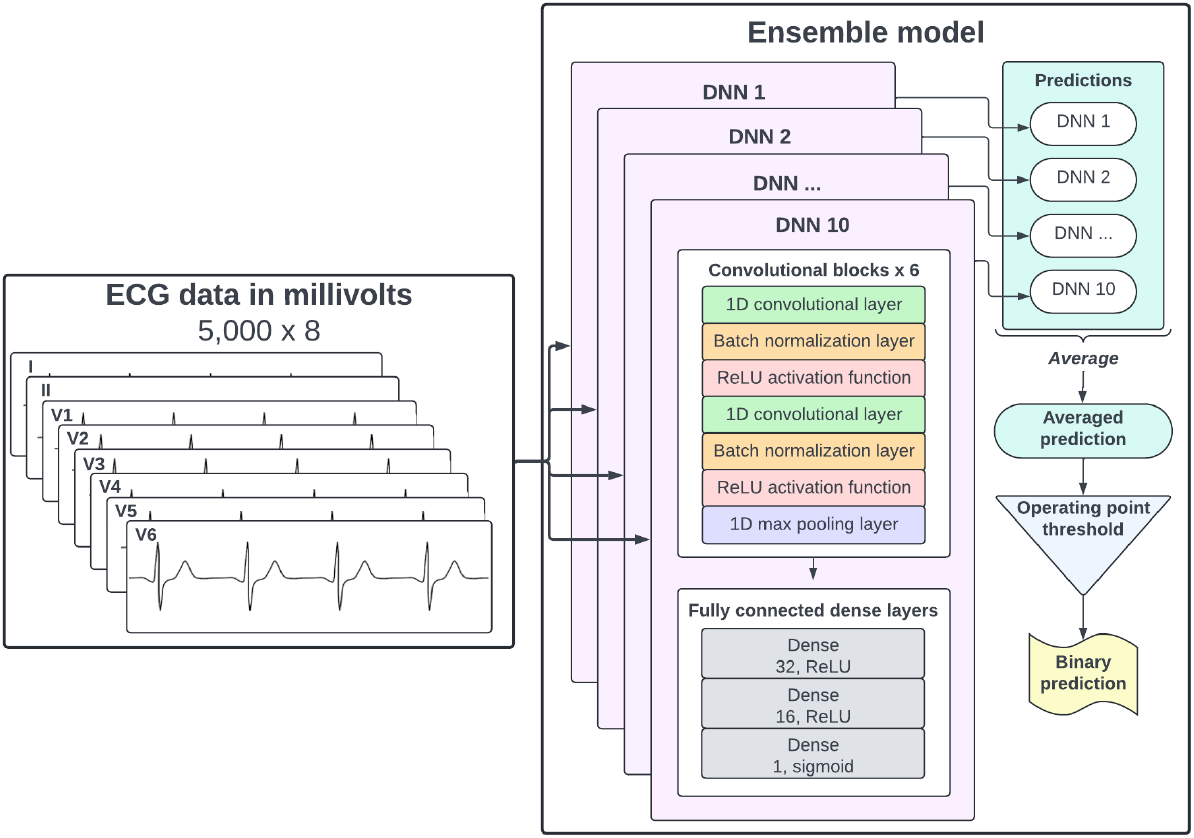
Model architecture.

## 2 Methods

### 2.1 Training Data

We assembled a multi-year dataset of paired 12-lead ECGs and transthoracic echos (TTEs) from nine hospitals and clinics for model training. ECGs were used as model inputs, and the EF value from the TTE served as the ground truth for each case, with low EF defined as *≤* 40%. The final training cohort consisted of 930,689 ECG-TTE pairs. We ran extensive experiments and ablation studies to optimize data selection, testing different prevalence rates, time windows around TTE, and the inclusion of paced and noisy ECGs. Results showed that the best outcome was achieved by including all ECGs from patients who never had low EF, and only ECGs taken within 30 days of a positive TTE for patients with low EF. Additionally, training with noisy ECGs improved robustness, but training with paced ECGs did not.

### 2.2 Model Architecture

During model development, we tested several model architectures, including transformers and convolutional neural networks (CNN). We found that an ensemble of 10 CNNs consistently performed best. Each CNN in the ensemble has the same architecture, differing only by random seed. Model inputs consist of 10-second, 8-lead ECG waveforms in millivolts. The architecture includes a convolutional feature extractor (convolutional layers, batch normalization, ReLU activations) followed by three fully connected layers and a final sigmoid output. Models were trained with the Adam optimizer, binary cross-entropy loss, early stopping, and learning rate reduction on plateau. The ensemble output score is calculated by averaging the predicted probabilities from the 10 models. The final binary classification is obtained by applying an operating point threshold (OP) to this averaged score. Separate OPs are used for the following populations: females <65, females 65+, males <65, and males 65+. All OPs were chosen to minimize the difference between sensitivity and specificity.

### 2.3 External Validation

This was a retrospective observational cohort study that was designed to evaluate the performance of our model using real-world data from four geographically diverse health systems in the United States, including a community hospital, academic medical center, integrated delivery system, and unified health system. Importantly, none of the data or patients at the external validation sites were used to develop the model. Power analysis using a one-sample proportion Z-test determined that a minimum sample size of 16,000 cases was required to achieve 80% power for sensitivity and 90% power for specificity. The power calculations used lower-bound estimates of sensitivity (83%) and specificity (84%) based on cross-validation experiments. Each of the four sites contributed at least 4,000 patients with linked resting ECG–TTE pairs. Eligible patients were required to be at least 40 years old at the time of ECG acquisition, to be at risk of heart failure, and to have at least one ECG–TTE pair, with the ECG performed within 30 days prior to the TTE. To ensure demographic diversity, pairs were selected in reverse chronological order and stratified by race and ethnicity to reflect the US census population.

The primary endpoints of the study were sensitivity and specificity with 95% Wilson confidence intervals. Both metrics were considered equally important for validating the model’s clinical utility. For the study to be considered successful, the lower bound of the confidence interval for both sensitivity and specificity was required to exceed 80%. Secondary endpoints included positive predictive value (PPV), negative predictive value (NPV), and diagnostic odds ratio (DOR).

**Figure 2.**
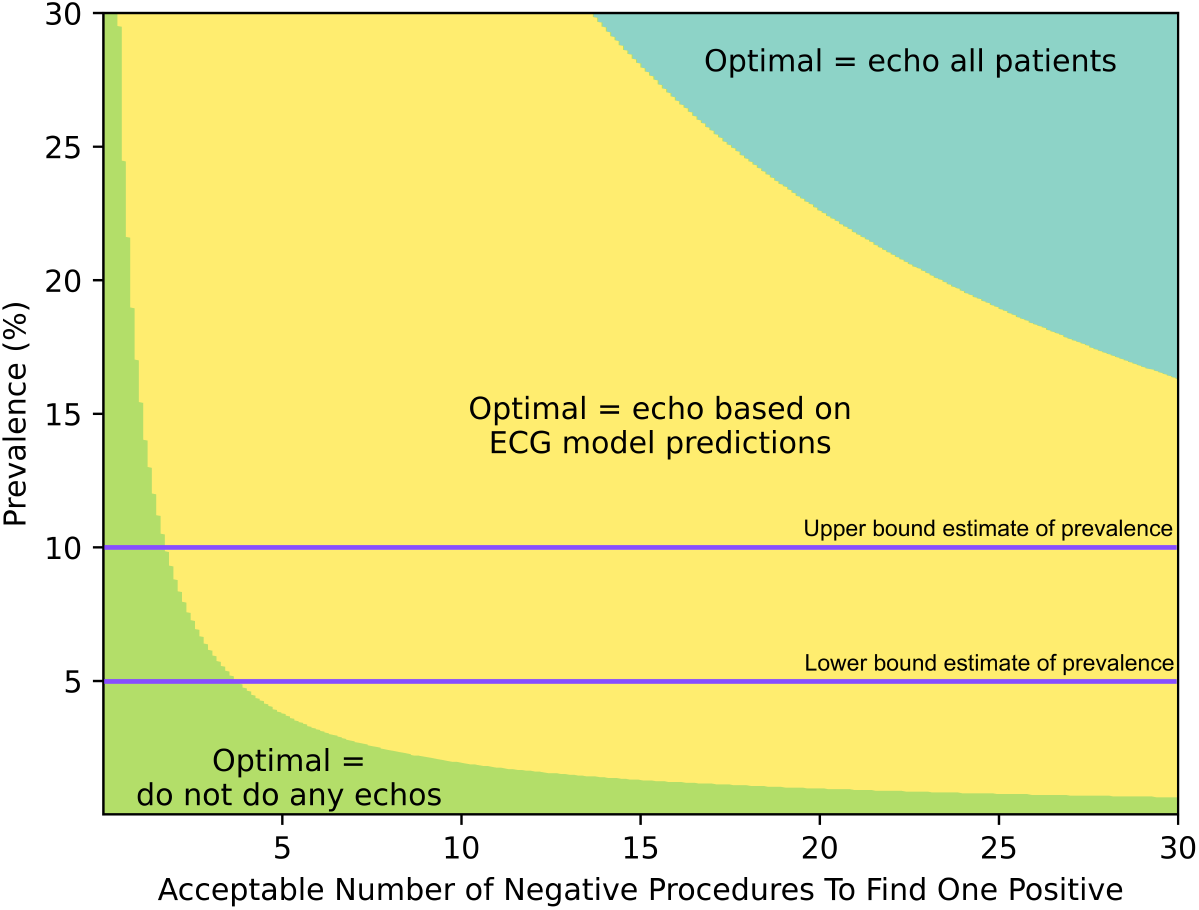
Clinical net benefit results. The figure compares the net benefit of three approaches: using the model to select patients for echo (yellow), echoing all patients (blue), and not performing any follow-up echos (green). The colors illustrate when each approach has the greatest net benefit as compared to the other approaches. Horizontal lines mark the lower and upper bound estimates of disease prevalence. When clinicians prefer to perform very few extra echos per true positive, or when disease prevalence is low, not performing additional echos yields the highest net benefit (green). Conversely, echoing all patients has the highest net benefit when disease prevalence is high and clinicians are willing to perform >14 follow-up echos to find one positive (blue). Everywhere else (yellow), the ECG model maximizes net benefit.

### 2.4 Clinical Net Benefit

To assess the clinical value of our model beyond metrics traditionally used in machine learning, we performed a clinical net benefit analysis (4). Net benefit is a measure that quantifies the trade-off between true positives (*TP*) and false positives (*FP*) at a given net benefit probability threshold (*p*_*t*_). Importantly, net benefit has the added advantage of being more interpretable to clinicians, as it allows them to incorporate their own expectations about disease prevalence and the number of acceptable procedures needed to find a positive case—without requiring specialized knowledge of machine learning or statistics. Net benefit is defined as follows:

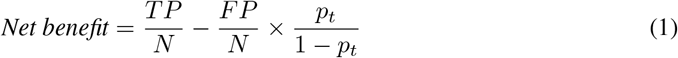

where *N* is the number of total samples, *TP* is true positives, *FP* is false positives, and *p*_*t*_ is the risk level at which a clinician would order further testing (i.e., echo). If *p*_*t*_ equals 10%, for example, any patient with a predicted risk of having low EF at or above 10% would be referred for follow-up.

Clinicians routinely apply thresholds like this in practice by weighing medical history, symptoms, and risk estimates to guide care. The threshold (*p*_*t*_) reflects the trade-off between detecting true cases and minimizing unnecessary follow-up, and it can vary based on clinician preferences and resource availability. To capture this variability, we calculated net benefit across a range of thresholds, each representing the number of follow-ups a clinician would be willing to perform to find one true case of low EF. This approach allows assessment of clinical net benefit under different, realistic decision scenarios. For additional context, we compared the model’s net benefit to the strategies of screening all patients or no patients; this provides a benchmark to ensure the model offers greater clinical value than simply referring everyone or no one for further testing. Given that clinicians cannot currently screen for low EF using only ECGs, the current standard of care is equivalent to screening no patients.

## 3 Results

### 3.1 External Validation

A total of 15,125 patients met the eligibility criteria for inclusion in the validation study. Of these, 201 were marked as unclassifiable due to poor quality and were excluded from the analysis set. This left us with a total of 14,924 ECG-TTE pairs (49% female; >25% from racial and ethnic minority groups). Low EF prevalence in this dataset was 10.7% overall. The median age was 70 years, with good representation across both younger (35% aged 40–64) and older (65% aged 65+) patient populations.

For the primary endpoints, the device achieved a sensitivity of 85.8% (lower bound: 84.0%) and a specificity of 83.0% (lower bound: 82.4%), both exceeding the prespecified acceptance criterion of 80%. Secondary endpoints included a PPV of 37.7%, NPV of 98.0%, and DOR of 29.49. Subgroup analyses showed that the device performed consistently across age, sex, race, ethnicity, clinical site, and ECG manufacturer. These results demonstrate robust clinical performance and support the device’s value for identifying patients with low EF.

### 3.2 Clinical Net Benefit

The number of follow-up procedures needed to find one true positive depends on the prevalence of low EF, which varies by clinical setting and is often underestimated due to underdiagnosis. In populations with clinically acquired ECGs, studies estimate the prevalence of undiagnosed low EF to be between 5% and 10% (5). The net benefit analysis showed that if a provider is willing to order five echos to find one new case, using the model is beneficial in a population with a 5% prevalence. However, if the provider is only willing to order three echos per positive case, the model should be used in a population with at least a 10% prevalence. The net benefit of the device is also influenced by the operating point (OP). For example, using a lower sensitivity OP would reduce the number of follow-ups needed per new case. Future work could optimize the OP to better match clinician preferences. Overall, these results demonstrate that the model provides meaningful clinical value in realistic screening scenarios. By quantifying this trade-off, net benefit analysis can help guide practical decisions about model adoption in diverse healthcare settings.

## 4 Conclusions and Future Work

Our model achieved high sensitivity and specificity for detecting low EF from resting 12-lead ECGs, with robust performance across demographic and clinical subgroups. Importantly, our use of clinical net benefit analysis moves beyond traditional metrics to quantify the real-world value of the model, showing that it provides tangible benefit over baseline strategies in typical clinical scenarios. With FDA clearance, this model can now be integrated into hospital systems, allowing us to observe how clinicians use the tool in practice and to measure its real-world impact on patient care and healthcare delivery. Future work will focus on understanding clinician adoption, workflow integration, and the downstream effects of model-guided screening on clinical outcomes and resource utilization.

## Data Availability

Deidentified data used in the research were collected in a real-world health care setting and are subject to controlled access for privacy and proprietary reasons. When possible, derived data supporting the findings of this study have been made available within the paper and its Supplementary Figures/Tables.

